# Fetal brain growth and infant autistic traits

**DOI:** 10.1101/2021.10.15.21264826

**Authors:** Ezra Aydin, Alex Tsompanidis, Daren Chaplin, Rebecca Hawkes, Carrie Allison, Gerald Hackett, Topun Austin, Eglė Padaigaitė, Lidia V. Gabis, John Sucking, Rosemary Holt, Simon Baron-Cohen

**Affiliations:** Autism Research Centre, Department of Psychiatry, University of Cambridge, UK; Department of Psychology, University of Cambridge, UK; The Rosie Hospital, Cambridge University Hospitals Foundation Trust, Cambridge, UK; NIHR Cambridge Biomedical Research Centre, Cambridge, UK; Wolfson Centre for Young People’s Mental Health, Cardiff University, UK; Sackler School of Medicine at Te Aviv University and Child Development Centre, Sheba Hospital, Israel; Department of Psychiatry, University of Cambridge, UK

**Keywords:** Early brain development, ultrasound, transcerebellar diameter, Q-CHAT, autistic traits

## Abstract

**Background:** Research indicates that structural differences exist in the brains of individuals who later display developmental conditions (e.g., autism). To date only a handful of studies have explored the relationship between fetal brain growth and later infant outcomes, with a particular focus on fetal head circumference (HC) as a proxy for brain development. These findings have been inconsistent. We investigate whether fetal brain measurements correlate with the emergence of autistic traits in toddlers.

**Method:** 219 singleton pregnancies (104 males and 115 females) were recruited at the Rosie Hospital, Cambridge, UK. A 2D ultrasound was performed at 12-, 20- and between 26-30-weeks of pregnancy, measuring head circumference (HC), ventricular atrium (VA) and transcerebellar diameter (TCD). 178 infants were subsequently followed up at 18-20 months of age and completed the Quantitative Checklist for Autism in Toddlers (Q-CHAT) to observe early autistic traits.

**Results:** HC was larger in males than in females in both the second and third trimester. There was a significant positive association between TCD size at 20 weeks and Q-CHAT scores at 18-20 months of age, found in both univariate and multivariate analyses, and this remained significant after controlling for sex.

**Conclusion:** There is a positive relationship between cerebellar (TCD) development at 20 weeks’ gestation and the later emergence of autistic traits (at 18-20 months of age). Atypical neurodevelopment may start prenatally. If replicated these findings could facilities early diagnosis and improved outcomes.

## Introduction

Approximately four weeks after conception, the formation of the cerebral hemispheres can be observed (Muller & O’Rahilly, 1988), with spontaneous movements of the head and trunk being seen from as early as 9 weeks’ gestation age (GA) (de Vries, Visser, & Prechtl, 1982). By 37-42 weeks’ GA, the fetus may be able to demonstrate a level of cognitive capacity, such as the ability to discriminate between sounds (Kadic & Kurjak, 2018) and to anticipate directed movements (Reissland, Francis, Aydin, Mason, & Schaal, 2014). Thus, during these first 40 weeks of gestation the foundations for later cognition and behaviour are programmed, likely by both genetic and environmental factors including prenatal hormonal influences and their interaction.

Longitudinal research observing the relationship between fetal growth and development in relation to later outcomes is still in its infancy. Currently, fetal biometrics are used to predict newborn infants who may be at risk for low birth weight due to intrauterine growth restriction (IUGR) or are small for gestational age as well as those who are large for gestation age (LGA). These risks have been identified as common predictors of adverse fetal and infant outcomes. Numerous studies have explored the relationship between deviation in fetal growth (e.g., LGA) and birth weight with mixed findings for later child outcomes (e.g. increased likelihood of developing sensory issues (Machado, Oliveira, Magalhães, Miranda, & Bouzada, 2017), autism (Beranova et al., 2017; Pyhälä et al., 2014) and increased incidence of cognitive deficits (Kok, Lya den Ouden, Verloove-Vanhorick, & Brand, 1998)). However, the majority of research examining the relationship between the prenatal period and later outcomes has only measured pregnancy outcomes (e.g., birthweight) with very few studies measuring the longitudinal relationship between fetal growth and development on early infant outcomes.

Heritability estimates for autism reveal a significant genetic component (Bai et al., 2019) which could affect brain development. The genetics of autism have been shown to overlap with genetic variance that is associated with baseline sex differences in growth and anthropometric measures (Mitra et al., 2016). Early brain overgrowth is a prominent theory in autism aetiology research (Constantino, Zhang, Frazier, Abbacchi, & Law, 2010; Dinstein, Haar, Atsmon, & Schtaerman, 2017; Hazlett et al., 2011; MRC AIMS Consortium et al., 2020) with numerous researchers observing the relationship between early (0-36 months) head growth and the later diagnosis of autism. However, there is a growing body of evidence demonstrating no differences in brain development in toddlers who are diagnosed with autism. Whilst Constantino et al (2010) reported a slight acceleration in head growth during the first 2 years of life, this margin of difference is not great enough to be considered a reliable marker of increased autism likelihood. Many studies (using HC as a proxy for brain growth) have observed no differences in longitudinal head growth from birth to infancy (Dinstein & Shelef, 2018; Zwaigenbaum et al., 2014), concluding that enlarged HC size is not reliably associated with autism (Dinstein et al., 2017) in postnatal development. However, this debate remains unresolved since the latest big data MRI study suggests there is increased brain volume and cortical thickness in children with autism (MRC AIMS Consortium et al., 2020).

The association between fetal growth, pregnancy outcomes and later autism likelihood is still relatively unclear (Bai et al., 2019). Studies exploring the relationship between prenatal brain growth and later infant outcomes have produced mixed findings. No significant difference on HC has been found between autistic children or siblings with an increased genetic likelihood for autism compared to non-autistic controls, in both prenatal and postnatal research (Blanken et al., 2018; Unwin et al., 2016; Whitehouse, Hickey, Stanley, Newnham, & Pennell, 2011). Unwin et al. (2016) suggested that examining the growth of subregions of the fetal brain may reveal potential differences between fetuses who later receive an autism diagnosis compared to those who do not, rather than using a proxy measure of brain growth (i.e., HC). Growth velocity in HC (both pre- and postnatally) has also been examined in relation to later developmental outcomes including autism (Regev et al., 2021). Abel et al., (2013) and Bonnet-Brilhault et al., (2018) both observed atypical prenatal head growth trajectories, describing overgrowth in prenatal HC during the second and third trimester. Whilst HC has been commonly used by researchers as a proxy for brain growth, studies have yet to investigate subregions of the developing fetal brain in relation to the later emergence of autism.

The cerebellum is one of the earliest structures to develop, emerging from the roof of the rhombencephalon between 4-6 weeks post-conception (Garel, Fallet-Bianco, & Guibaud, 2011) putting it at increased vulnerability to a range of developmental effects during prenatal development (Garel et al., 2011; Koning et al., 2017). Transcerebellar diameter (TCD) in fetuses measuring below the 5^th^ centile is a relevant marker to detect associated anomalies such as a high rate of fetal malformations, chromosomal anomalies, severe IUGR and genetic disorders (Atallah et al., 2019). Reduced fetal cerebellar diameter has been implicated in atypical general movement in infants at 1 and 3 months old (Spittle et al., 2010) and poor motor and cognitive functions (Park et al., 2014). Similarly, studies have shown that prenatal isolated ventriculomegaly is associated with later developmental delay (e.g., in fine motor and expressive language skills (Lyall et al., 2012)) and psychiatric diagnoses (autism and schizophrenia) (Gilmore et al., 1998; Gilmore et al., 2001; Palmen et al., 2005).

The aim of this study was to observe growth of the cerebrum (using HC as a proxy) (Chang, Chang, Yu, Ko, & Chen, 2000), the cerebellum (Rutten et al., 2009) and the ventricular atrium (VA) (Achiron et al., 2004; Kim, Jeanty, Turner, & Benoit, 2008) of fetuses measured with 2D ultrasound sonography in the 2^nd^ and 3^rd^ trimesters of gestation. Subsequently, infants were followed up and a potential association was investigated between these regions of the developing fetal brain and the early emergence of autistic traits (Q-CHAT scores) at 18-20 months of age.

## Methods

### Participants

219 healthy fetuses (104 males and 115 females) from neurotypical mothers were recruited prospectively from the Rosie Maternity Hospital in Cambridge, UK. Inclusion criteria for the study were as follows: pregnant women who were willing to have an additional ultrasound scan between 26-30 weeks’ gestation (average GA:28 weeks, SD=1.25), with (1) little/no consumption of alcohol during pregnancy, (2) no smoking or recreational drug use during pregnancy, (3) a singleton fetus whose measurements indicated their size to be appropriate for GA and (4) the absence of any major fetal anomalies. For inclusion in the final data analysis, the birth resulted in a clinically healthy baby (see T*able* 1 for characteristics of the mothers and fetuses).

### Ethics

A favourable ethical opinion for the study was given by the East of England Cambridge Central Research Ethics Committee (REC Ref:16/EE/0004). All mothers gave written informed consent for access to previous scans, additional scans and follow-ups.

### Procedure

Ultrasound scans were performed using a GE Voluson 8 Expert ultrasound system, with a (4-8 MHz curvilinear abdominal transducer). All women had completed a normal 12- and 20-week anomaly scan and were made aware that the additional scan (between 26-30 weeks’ gestation) was for research purposes and was not a routine medical scan. During various stages of fetal development, standard ultrasound measurements were taken (*Table 1*). Fetal brain measures included HC, VA and TCD. These measurements were taken by trained sonographers using standard ultrasound planes specific to each fetal measurement. HC was measured with a standard clinical protocol by obtaining a cross-sectional view of the fetal head at the level of the ventricles and measuring around the outer edge of the skull. With this view, the posterior atrium of the VA was also measured (*Figure 1*). For TCD, the back of the fetal head was visualised keeping in view the septum pellucidum. To measure the diameter, electronic callipers were placed on the outer, lateral edges of the cerebellum (*Figure 2*). The research team were given consented access to the mothers’ previous 12- and 20-week routine medical ultrasound scans and HC, VA and TCD obtained as described above.

**Table 1:**
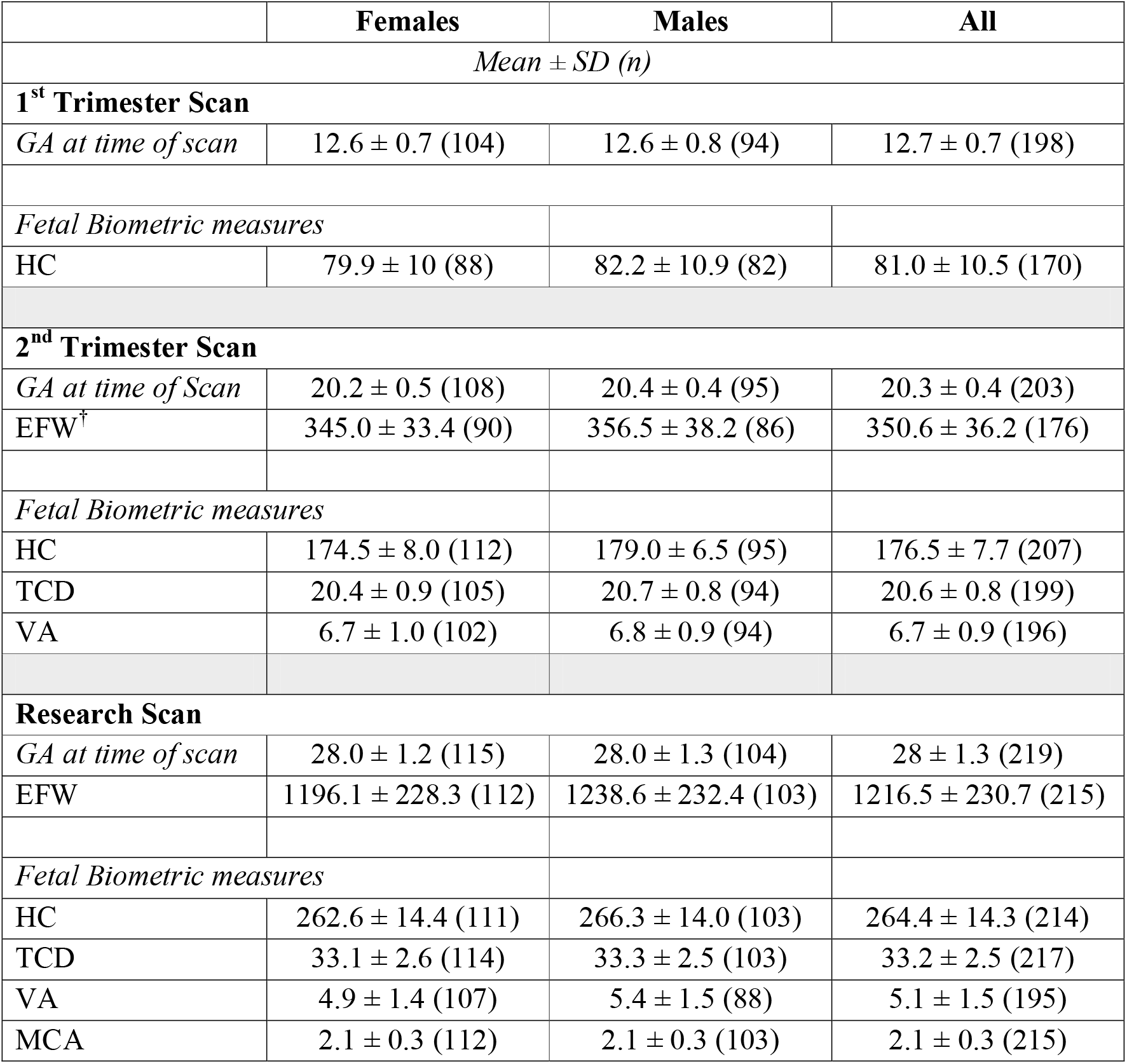
Characteristics of fetal biometrics in the study, separated by sex.

|  | Females | Males | All |
| --- | --- | --- | --- |
| *Mean ± SD (n)* |  |  |  |
| **1<sup>st</sup> Trimester Scan** |  |  |  |
| *GA at time of scan* | 12.6 ± 0.7 (104) | 12.6 ± 0.8 (94) | 12.7 ± 0.7 (198) |
| *Fetal Biometric measures* |  |  |  |
| HC | 79.9 ± 10 (88) | 82.2 ± 10.9 (82) | 81.0 ± 10.5 (170) |
| **2<sup>nd</sup> Trimester Scan** |  |  |  |
| *GA at time of Scan* | 20.2 ± 0.5 (108) | 20.4 ± 0.4 (95) | 20.3 ± 0.4 (203) |
| EFW<sup>†</sup> | 345.0 ± 33.4 (90) | 356.5 ± 38.2 (86) | 350.6 ± 36.2 (176) |
| *Fetal Biometric measures* |  |  |  |
| HC | 174.5 ± 8.0 (112) | 179.0 ± 6.5 (95) | 176.5 ± 7.7 (207) |
| TCD | 20.4 ± 0.9 (105) | 20.7 ± 0.8 (94) | 20.6 ± 0.8 (199) |
| VA | 6.7 ± 1.0 (102) | 6.8 ± 0.9 (94) | 6.7 ± 0.9 (196) |
| **Research Scan** |  |  |  |
| *GA at time of scan* | 28.0 ± 1.2 (115) | 28.0 ± 1.3 (104) | 28 ± 1.3 (219) |
| EFW | 1196.1 ± 228.3 (112) | 1238.6 ± 232.4 (103) | 1216.5 ± 230.7 (215) |
| *Fetal Biometric measures* |  |  |  |
| HC | 262.6 ± 14.4 (111) | 266.3 ± 14.0 (103) | 264.4 ± 14.3 (214) |
| TCD | 33.1 ± 2.6 (114) | 33.3 ± 2.5 (103) | 33.2 ± 2.5 (217) |
| VA | 4.9 ± 1.4 (107) | 5.4 ± 1.5 (88) | 5.1 ± 1.5 (195) |
| MCA | 2.1 ± 0.3 (112) | 2.1 ± 0.3 (103) | 2.1 ± 0.3 (215) |

**Figure 1:**
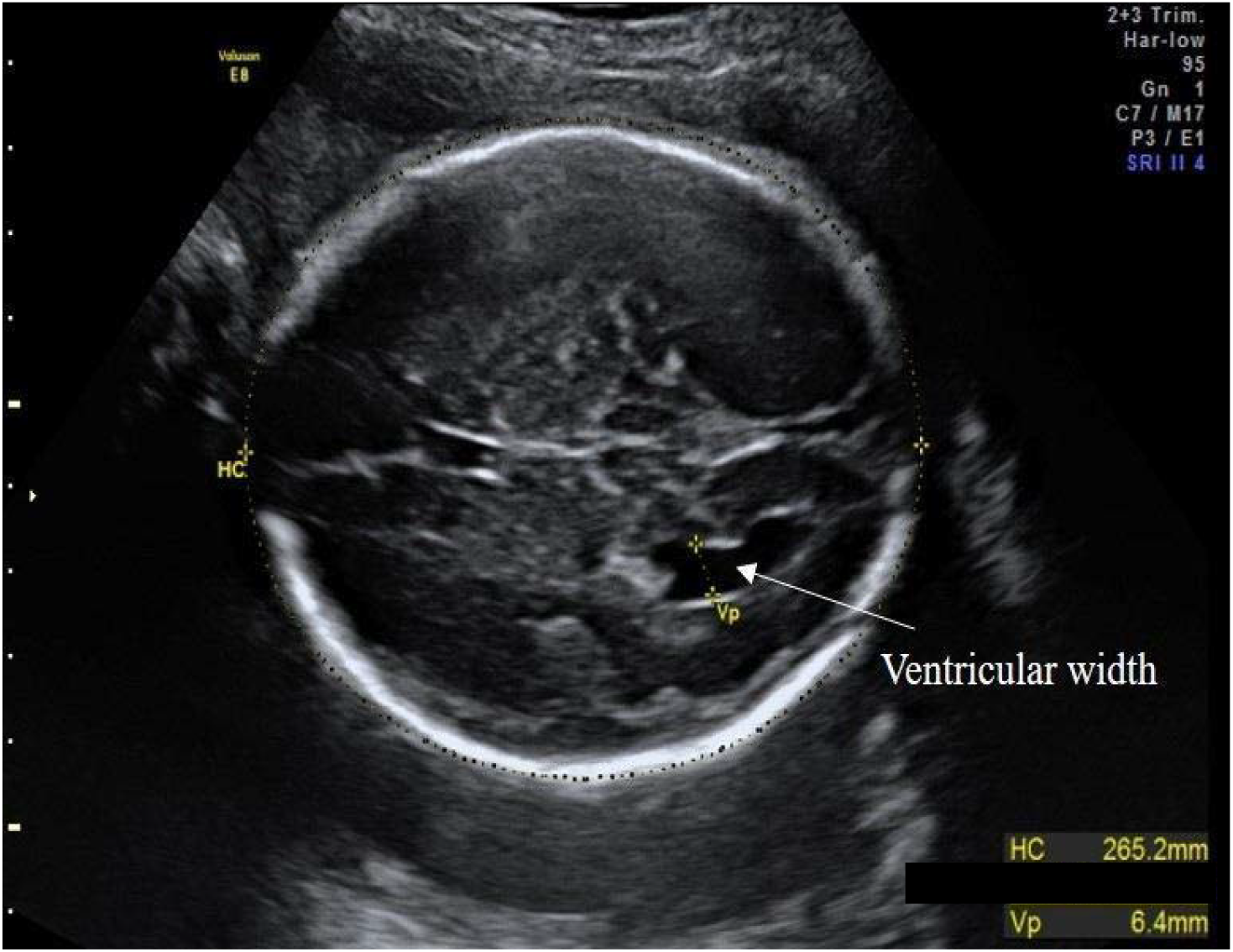
Example of a posterior VA measurement in a female fetus. VA was measured slightly above the level of the thalami in the axial plane of the fetal brain. Electronic callipers were placed perpendicular to the long axis of the ventricle (highlighted by the yellow dotted line).

**Figure 2:**
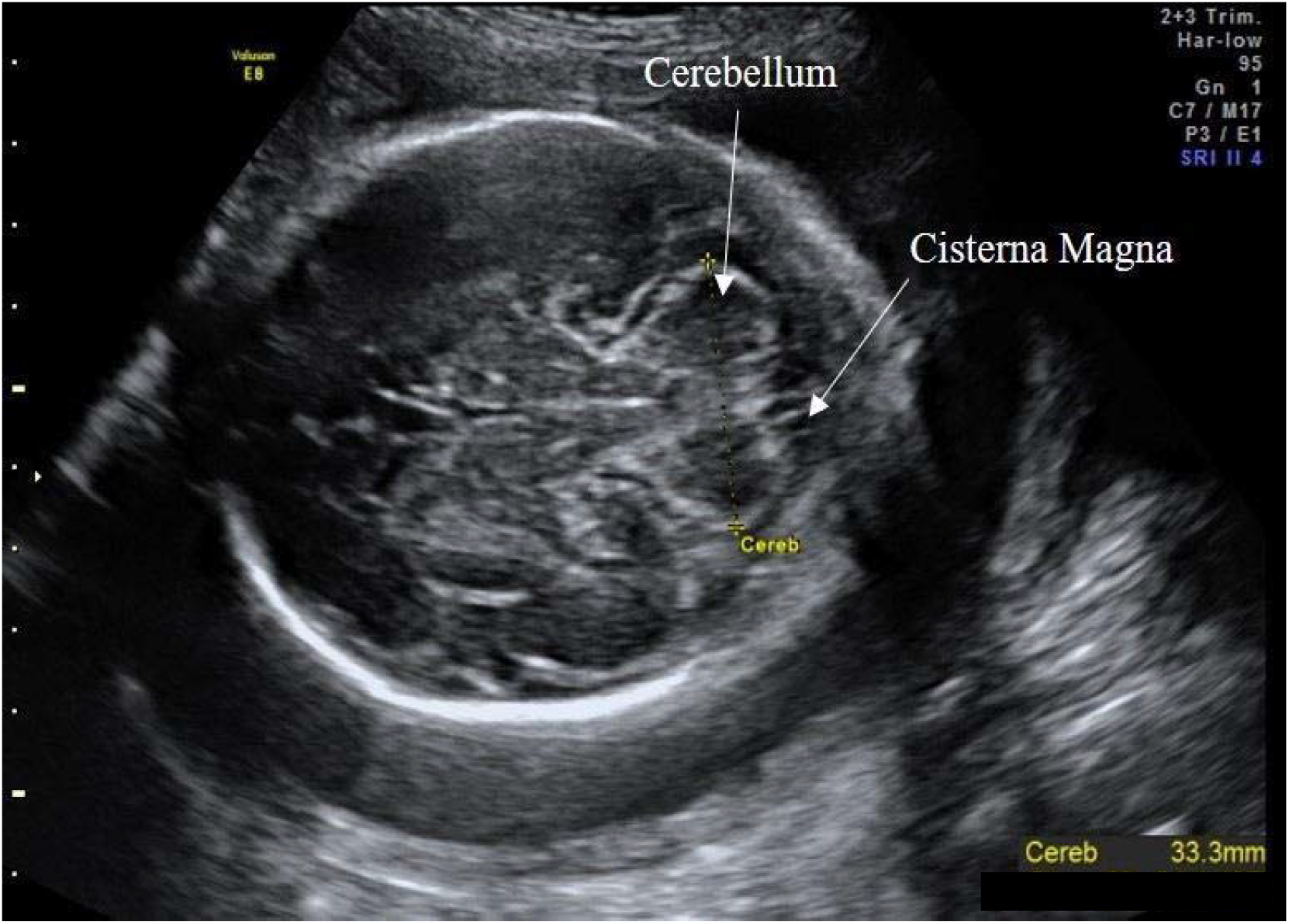
Example of the TCD measurement in a female fetus. For TCD measurements, the transducer was slightly rotated from the thalamic plane (used to measure HC) until the posterior fossa was visible. TCD was measured using the ‘outer-to-outer’ method. TCD taken during the research scan always showed LGA. This is due to the fact that the internal growth charts on the GE Voluson 8 Expert ultrasound scanner (Hill, Guzick, Fries, Hixson, & Rivello, 1990) did not cover this GA. However, all TCD were within expected 5-95 centile ranges.

If any of the measurements were not feasible due to fetal positioning the mother was asked to walk around for several minutes or asked to come for a second appointment. In instances where the fetus was breech in presentation, the examination bed was tilted to move the fetus out of the pelvis and remove shadowing whilst the measurement was taken. Inability to take a measurement during scanning was mainly a result of fetal positioning and/or high maternal BMI.

### 18 month-follow up

When the infants were between 18-20 months’ (mean age; 18 months’ and 22 days), parents were sent an online version of Quantitative Checklist for Autism in Toddlers (Q-CHAT, validated in ages 18-30 months (Allison et al., 2021)). 178 parents completed this online follow-up; 92 females (mean:29.6, SD:7.6), 85 males (mean:30.0, SD:7.8).

### Statistics

#### Analysis

All statistical testing was performed in RStudio. Q-CHAT scores were assessed for skewness and kurtosis and for outliers. Extreme outliers were reduced to the maximum value within the range defined by the 1^st^ and 3^rd^ quartiles plus 1.5 times the interquartile range. Possible independent predictors of Q-CHAT score, including cohort covariates (*Table 2*), were initially assessed via univariate regression by calculating the Pearson’s correlation coefficient. Binary categorical variables of the cohort were assessed via student’s t-tests. Ultrasound measures (dependent variable) were regressed on Q-CHAT scores, (independent variable) in separate univariate linear regression models (Model 1). Ultrasound measures were standardised to GA at the time of measurement using z-scores, with the adjusted measure reassessed for association with Q-CHAT score via univariate linear regression (Model 2). Multiple linear regression models for each ultrasound parameter were used to control for confounding effects of sex, maternal age and birth weight (Model 3).

**Table 2:** Association of brain parameters with Q-CHAT score: Model 1: Univariate linear regression to Q-CHAT score. Effect size uses Pearson’s correlation coefficient. Model 2: Univariate linear regression to Q-CHAT score with independent variable first standardised to GA at the time of measurement. Effect size uses Pearson’s correlation coefficient. Model 3: Multiple regression model of brain ultrasound parameter to Q-CHAT score, with infant sex, maternal age and birth weight as added covariates. Effect size is the semi-partial correlation coefficient of the specific ultrasound parameter in the model. R^2^ refers to the variance captured by the entire model.

|  | Weeks of Gestation |  |  |  |  |  |
| --- | --- | --- | --- | --- | --- | --- |
| Head Circumference |  |  |  |  |  |  |
|  | 12 Weeks |  | 20 Weeks |  | 28 Weeks |  |
|  | *Effect size* | *p* | *Effect size* | *p* | *Effect size* | *p* |
| Model 1 | r=0.008 | p=0.928 | r=0.105 | p=0.172 | **r=0.189** | p=**0.012** |
| Model 2<br>(Adjusted) | r=0.135 | p=0.116 | r=0.069 | p=0.374 | **r=0.16** | p=**0.035** |
| Model 3<br>(Multiple) | r=0.01 | p=0.929 | **r=0.18** | **p=0.022** | r=0.15 | p=0.058 |
|  | Model 3 R<sup>2</sup>= 0.018 |  | Model 3 R<sup>2</sup>=0.05 |  | Model 3 R<sup>2</sup>=0.046 |  |
| Transcerebellar Diameter |  |  |  |  |  |  |
|  | 12 Weeks |  | 20 Weeks |  | 28 Weeks |  |
|  |  |  | *Effect size* | *p* | *Effect size* | *p* |
| Model 1 |  |  | **r=0.233** | **p=0.003** | r=0.141 | p=0.060 |
| Model 2<br>(Adjusted) |  |  | **r=0.186** | **p=0.018** | r=0.13 | p=0.081 |
| Model 3<br>(Multiple) |  |  | **r=0.240** | **p=0.003** | r=0.13 | p=0.107 |
|  |  |  | Model 3 R<sup>2</sup>=0.073 |  | Model 3 R<sup>2</sup>=0.009 |  |
| Ventricular Atrium |  |  |  |  |  |  |
|  | 12 Weeks |  | 20 Weeks |  | 28 Weeks |  |
| Model 1 |  |  | β=0.093 | p=0.242 | r=-0.02 | p=0.796 |
| Model 2<br>(Adjusted) |  |  | β=0.097 | p=0.225 | r=-0.03 | p=0.682 |
| Model 3<br>(Multiple) |  |  | r=0.07 | p=0.414 | r=-0.06 | p=0.455 |
|  |  |  | Model 3 R<sup>2</sup>=0.022 |  | Model 3 R<sup>2</sup>=0.045 |  |

## Results

Sex differences in brain parameters were assessed in a total of n=219 fetuses (104 males, 115 females). Q-CHAT data were available for 178 toddlers (92 females and 86 males).

### Brain ultrasound measurements

Brain ultrasound measures correlated to GA to varying degrees at most time-points (*Supplemental Table 1, Supplemental Figure 1*). Measurements of HC and TCD showed a linear pattern of growth, over three and two consecutive time-points respectively (*Supplemental Figure 2*). There was no significant difference in the rate of growth of HC between the three measurements. VA did not significantly change between the second and third trimester and only correlated with GA at the third trimester. HC and TCD were significantly correlated with each other, at both the second and third trimester (2^nd^ trimester: Pearson’s r = 0.56, p<0.00001, 3^rd^ trimester: Pearson’s r = 0.824, p<0.00001), as well as to GA at the time of measurements (*Supplemental Figures 2 and 3*)

### Sexual dimorphism

There were no sex differences in measurements taken during the first trimester. HC was significantly larger in males, in the second trimester (*t* (204.7) = -4.53, *p* = 0.00001) (*Supplemental Figure 2*), but this was less apparent in the third trimester (*t* (211.4) = -1.94, *p* = 0.05). VA was larger in males in the third trimester (*t* (176.2) = -2.6, *p* = 0.01) but not the second (*t* (194) = -1.09, *p* = 0.28). TCD was larger in males in the second trimester, (*t (*196.9) = -1.96, *p* = 0.05), but this was not seen in the third trimester (*t* (202.5) = -0.77, *p* = 0.45).

### Q-CHAT Scores

A child’s age at the time of Q-CHAT scoring spanned a narrow range (mean=569.95 days, SD=21.68 days). The mean score (n=178) was 30.31 (SD=8.11). Distribution of scores showed a skew towards lower scores. One extreme outlier was reduced via winsorizing, to facilitate subsequent linear regression tests. There was no significant difference on Q-CHAT scores between males and females (*t*(172.6)=0.55, *p*=0.58). Other maternal parameters, such as a family history of autism, age, BMI, and parity did not significantly predict Q-CHAT score (*Supplementary Table 1 and 2*).

HC showed a borderline correlation with Q-CHAT score only after fetal sex and maternal age were included as covariates in multiple regression (Model 3) (*Table 2*). No significant associations were identified at 20- or between 26-30 weeks’ gestation of VA with Q-CHAT score.

There was a significant positive correlation between TCD at 20 weeks’ gestation and Q-CHAT score (Pearson’s correlation: r=0.23, p=0.003). This remained after standardising TCD measurements for GA (*Table 2*) and when controlling for fetal sex, birth weight and maternal age in a multiple regression model (Model 3). Interaction analysis also showed that the effect of TCD on Q-CHAT score was independent of sex (TCD: p=0.038) and that the interaction of TCD and sex did not significantly contribute to Q-CHAT score (interaction term: p=0.78). The same trend was noted for TCD at 28 weeks’ gestation, but this was no longer statistically significant in any of the models (Pearson’s correlation: r=0.14, p=0.06) (*Figure 3*).

**Figure 3:**
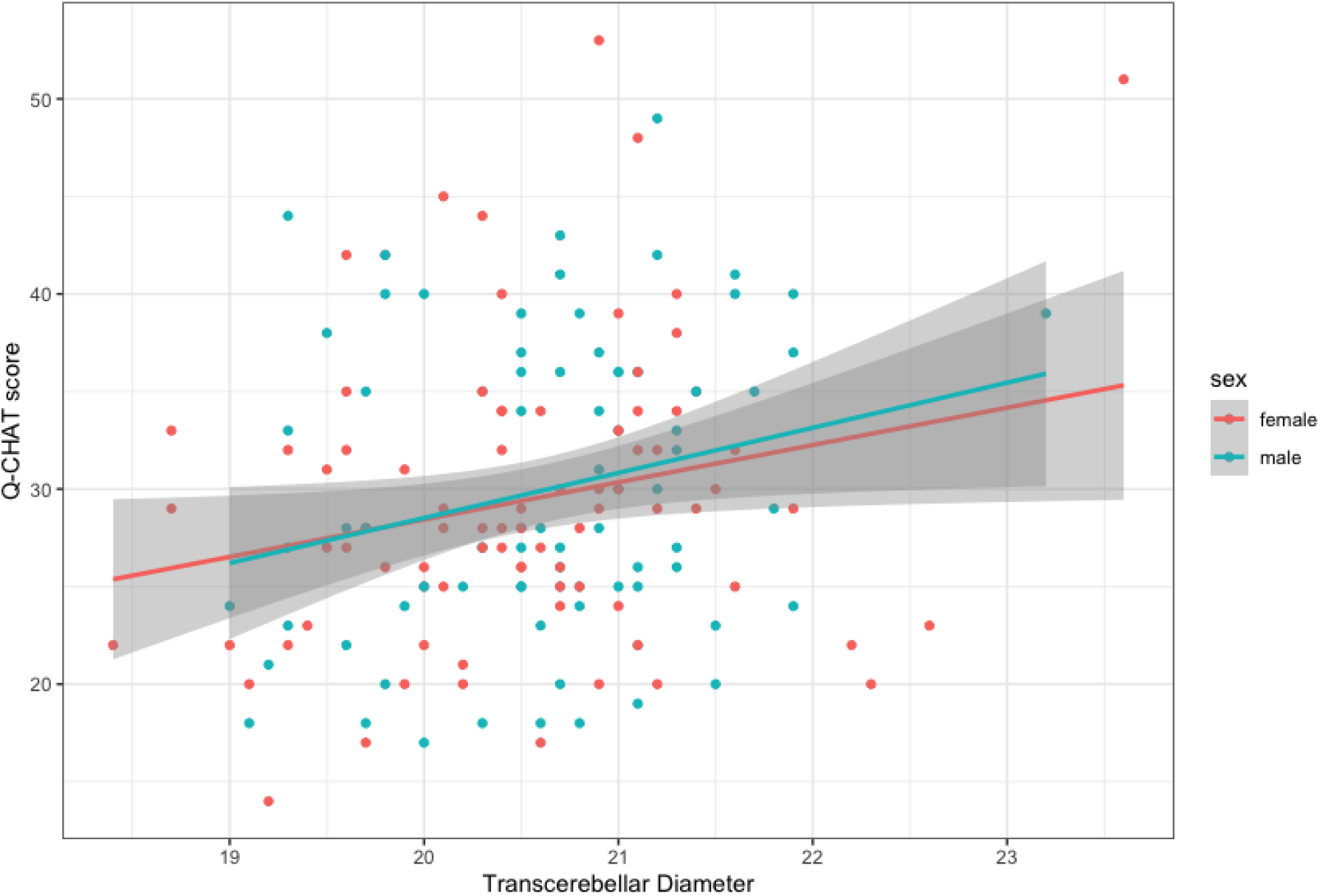
A scatterplot of TCD in the 2^nd^ trimester and Q-CHAT scores. Mean linear regression lines have been plotted by sex.

## Discussion

This is the first study to examine brain growth longitudinally *in utero* and associate it to early neurodevelopmental outcomes in infants. Specifically, we found that TCD in the 2^nd^ trimester was significantly associated with autistic traits measured by the Q-CHAT, at 18 to 20 months of age (*Figure 3*). Longitudinal ultrasound measures of fetal brain parameters showed varying rates of growth between trimesters and correlations between them (*Supplemental Figure 1*). HC and TCD were significantly correlated, particularly in the 3^rd^ trimester. HC increased at a stable rate from the 1^st^ to the 3^rd^ trimester, which was mirrored by increases in TCD between the 2^nd^ and 3^rd^ trimester *(Supplemental Figure 2)*. Sexual dimorphism was largely confined to the late 2^nd^ trimester, with a significant male advantage for both. This could be attributed to potential trophic effects from a sex steroid surge which occurs in male fetuses and peaks around the 16th week of gestation (Lombardo et al., 2012). A reduction of sex differences in the 3rd trimester is indicating potential “catching-up” for female fetuses before the end of the pregnancy, indicating that studies of postnatal brain parameters may not be accounting for prenatal sex differences.

TCD was correlated with Q-CHAT scores in the second trimester, after controlling for multiple covariates, but this was less apparent in the third. On the contrary, we found a significant effect of HC in the 3rd trimester but not in the 2nd (*Table 2*), The cerebellum contains more neuronal cell bodies than the neocortex and may thus be more responsive to trophic signalling earlier in pregnancy and prior to the completion of prenatal cortical expansion in the 3rd trimester (Herculano-Houzel, 2010). Fetal MRI could potentially reveal more microscopic increases in neuronal density at this developmental time-point and whether these are associated with later autistic traits.

The cerebellum has an important role in the regulation of neurodevelopment, as it regulates the structure and guides the formation of networks in the developing cortex (Ackerman, 1992). Early cerebellar dysfunction has been proposed to lead to autism, by preventing the integration of sensory stimuli and the maturation of the social association network in later life (Wang, Kloth, & Badura, 2014). This is consistent with analyses showing that genes associated with autism are more active during prenatal cerebellar development, when these cortical projections are established (Willsey et al., 2013).

Besides genetics, specific factors in the prenatal environment could be driving both TCD growth, as well as an increase in autistic traits. For example, oestradiol increases the density of neuronal fibres and spines in the developing cerebellum (Sasahara et al., 2007), as well as the likelihood of an autism diagnosis (Baron-Cohen et al., 2019). In addition, given the cerebellum’s rapid growth in late pregnancy, increases in TCD could be seen as an adaptive response to prenatal adverse conditions, which can also affect neurodevelopment (Clifton, 2010; Glynn & Sandman, 2011).

This cohort of pregnant mothers was relatively homogeneous in terms of clinical history and pregnancy parameters. Comorbidities such as polycystic ovary syndrome (PCOS), high BMI and age in the mother were not significantly associated with Q-CHAT score in the child. This does not necessarily contradict previously reported associations with autism (Cherskov et al., 2018; K. Lyall, Pauls, Santangelo, Spiegelman, & Ascherio, 2011), as this current study did not include clinically diagnosed cases and had a limited sample of 178 mother-child pairs with a Q-CHAT score. Similarly, there was no observed sex difference in autistic traits at 18-20 months of age, which could be attributed to a smaller sample size, compared with previous studies (Allison et al., 2008). Due to the nature and location of the study, ascertainment bias could also have contributed to the recruitment of volunteer mothers with a higher than typical range of autistic traits and/or with a significant interest in the neurodevelopment of their children.

In conclusion, this study demonstrated that prenatal brain growth is associated with early neurodevelopmental outcomes in infants. Specifically, we have shown that a larger TCD, as early as week 20, is associated with autistic traits in both males and females at 18-20 months. This finding further supports the potential inclusion of prenatal TCD as an early marker of later neurodevelopmental outcomes. The observed positive relationship between prenatal TCD size at 20 weeks’ gestation and Q-CHAT score at 18-20 months could be introduced into models of early postnatal screening for autism, with the aim of enabling early diagnosis and improving access to interventions to those that would benefit the most. However further longitudinal exploration is needed to validate this potential early marker, as well as its specificity to autistic traits. Finally, this finding may improve our understanding of how autism and related traits are associated with prenatal brain development.

## Key points

- Researchers have long debated the ‘early brain overgrowth’ theory and its relationship with autism.
- HC has been commonly used by researchers as a proxy for brain growth, with mixed findings. Studies have yet to investigate subregions of the developing fetal brain in relation to the later emergence of autism
- Our findings demonstrated a positive relationship between TCD size, at 20 weeks’ gestation autistic traits at 18 months of age.
- We identify the use of this standardised ultrasound measure as a potential early postnatal screen for autism, with further validation it has potential as a marker to guide improved access to early interventions to people that may benefit from it.

## Data Availability

All data produced in the present study are available upon reasonable request to the authors.

## Supplementary materials

**Suppl. Figure 1:**
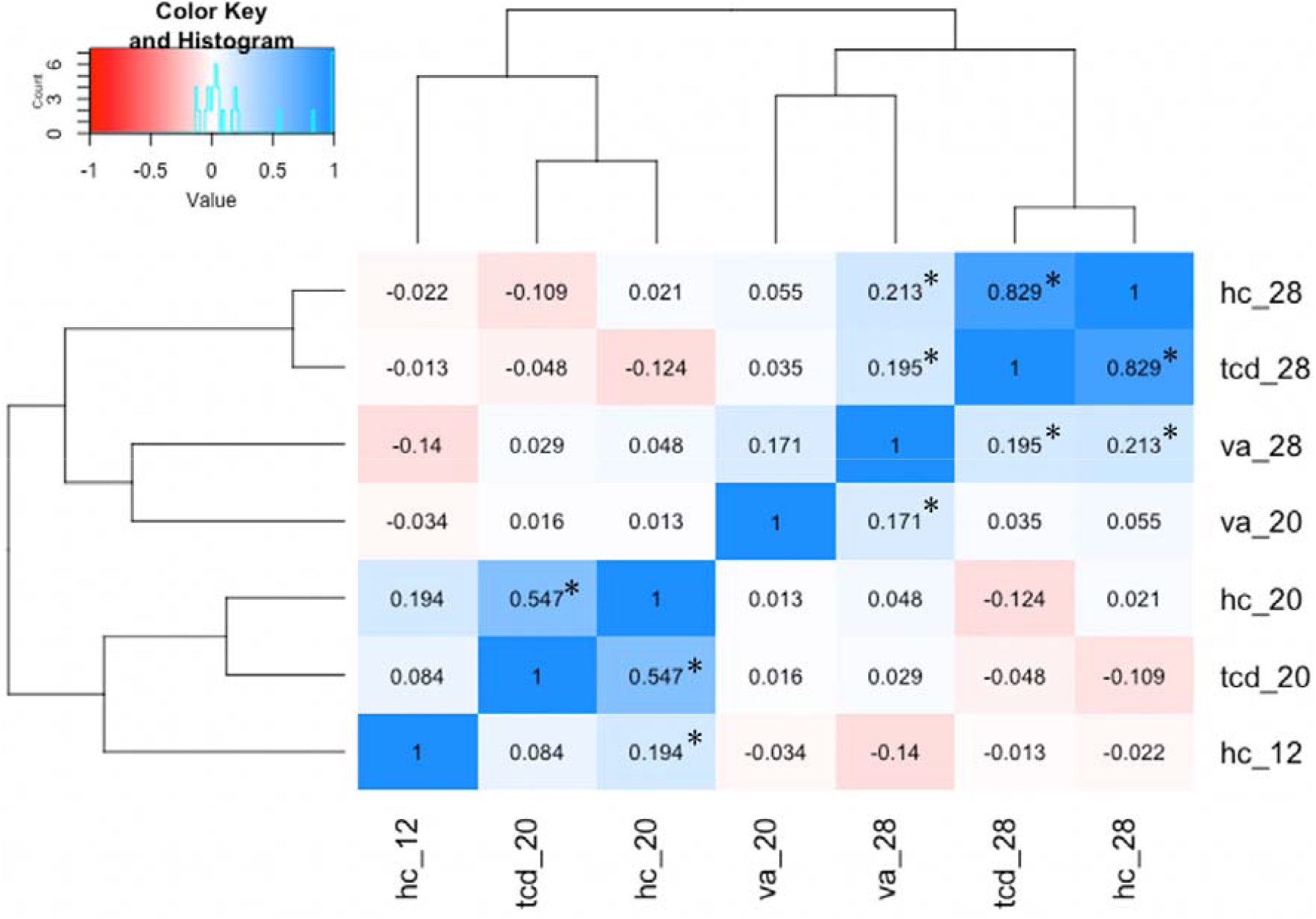
A heatmap and dendrogram with the pairwise associations of the measured brain parameters, at various time-points. Values indicate the Pearson’s correlation coefficient. The dendrograms along the top and left side show how the brain parameters and the rows are independently associated. *: p<0.05 hc_20/ 28: head circumference at 1^st^, 2^nd^ and 3^rd^ trimester respectively tcd_20/ 28: transcerebellar diameter at 2^nd^ and 3^rd^ trimester respectively va_20 / 28: ventricular atrium at 2^nd^ and 3^rd^ trimester respectivelyrespectively

**Suppl. Figure 2:**
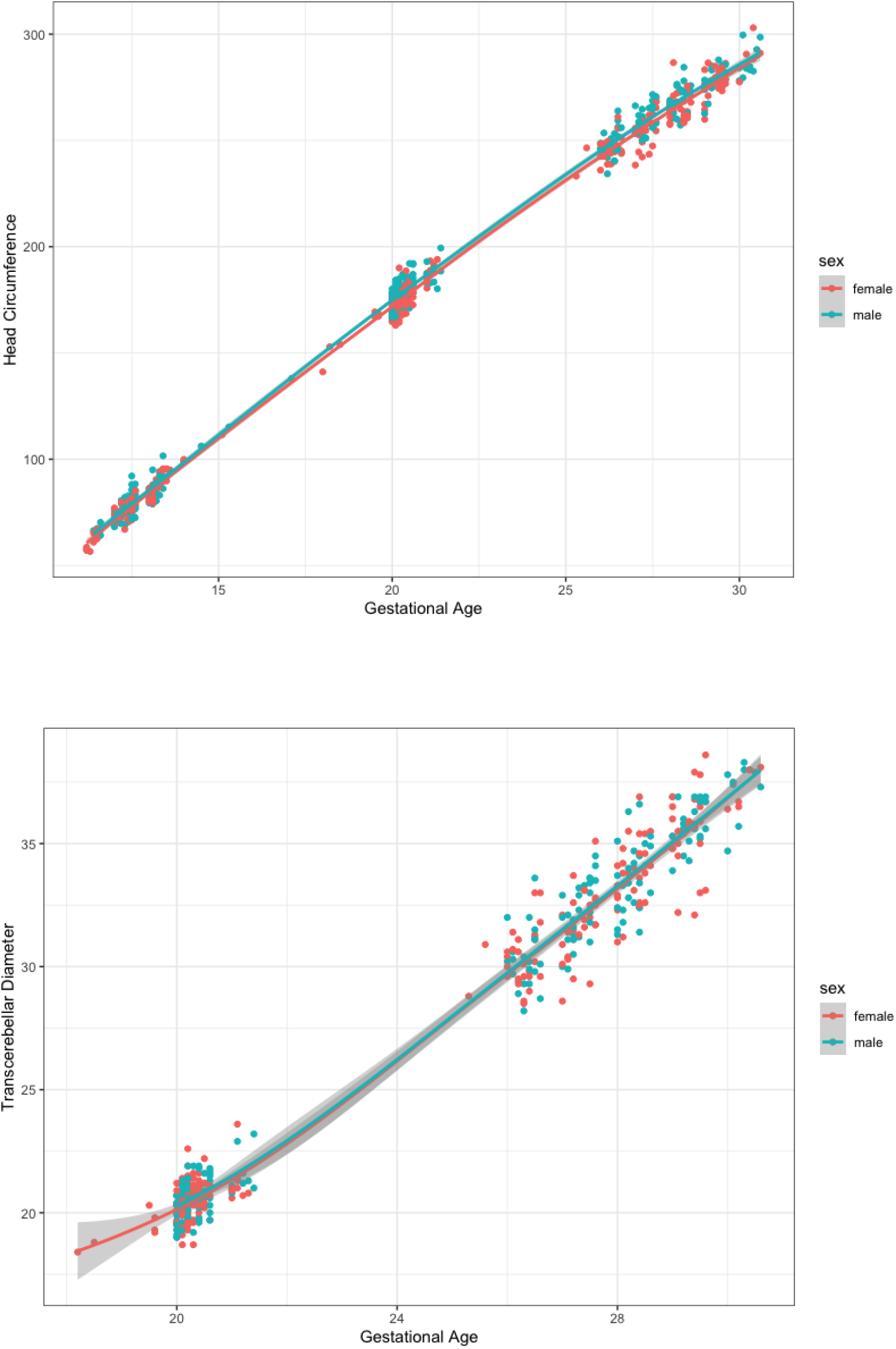
Ultrasound measurements plotted against GA at the point of assessment, with fitted curves of each sex for: A: head circumference, B: transcerebellar diameter

**Suppl. Table 1:** Pearson’s correlation coefficient of Q-CHAT scores with continuous maternal variables, showed no confounding effects.

|  | **Cohort**<br>**characteristics**<br>**Mean ± SD** | **Q-CHAT**<br>**correlation:**<br>**Pearson's $\beta$** | **Q-CHAT**<br>**correlation:**<br>**p-value** |
| --- | --- | --- | --- |
| **Infant age at Q-CHAT (in days)** | 570 ± 21.7 | -0.11 | 0.19 |
| **Maternal age (years)** | 32.4 ± 4.54 | -0.11 | 0.15 |
| **Maternal BMI (at 12-weeks' gestation)** | 26.3 ± 4.54 | 0.05 | 0.59 |
| **Parity** | 2.13 ± 1.27 | -0.08 | 0.3 |

**Suppl. Table 2:**
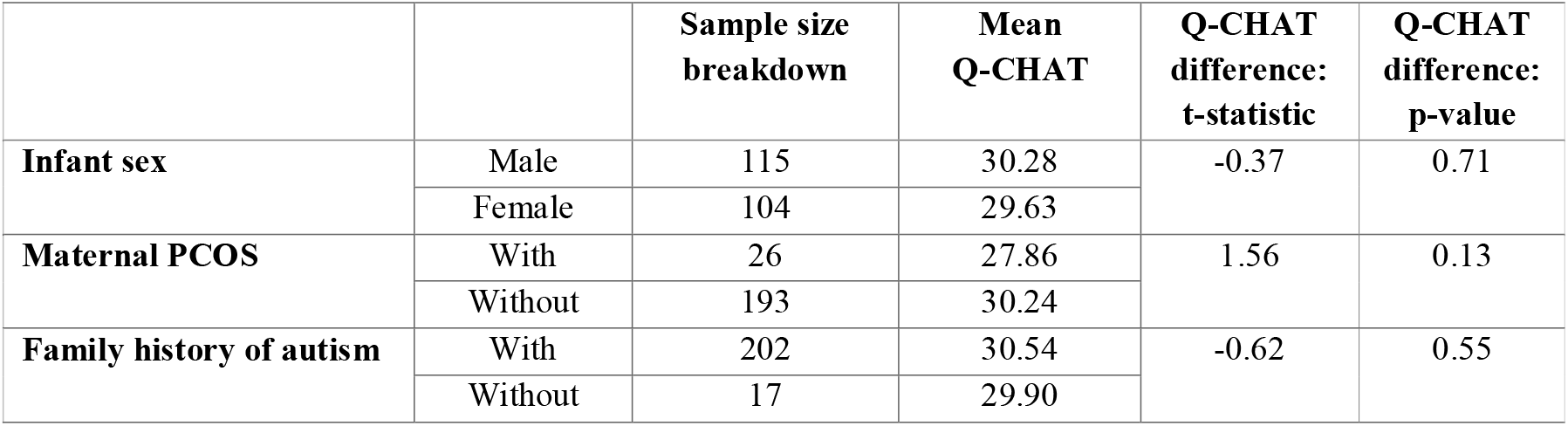
Q-CHAT score did not significantly differ between groups of potentially confounding categorical variables, when compared with two-tailed t-test.

|  |  | **Sample size**<br>**breakdown** | **Mean**<br>**Q-CHAT** | **Q-CHAT**<br>**difference:**<br>**t-statistic** | **Q-CHAT**<br>**difference:**<br>**p-value** |
| --- | --- | --- | --- | --- | --- |
| **Infant sex** | Male | 115 | 30.28 | -0.37 | 0.71 |
|  | Female | 104 | 29.63 |  |  |
| **Maternal PCOS** | With | 26 | 27.86 | 1.56 | 0.13 |
|  | Without | 193 | 30.24 |  |  |
| **Family history of autism** | With | 202 | 30.54 | -0.62 | 0.55 |
|  | Without | 17 | 29.90 |  |  |

